# *Chlamydia trachomatis* and *Neisseria gonorrhoeae* bacterial loads in co-infected men who have sex with men on pre-exposure prophylaxis: a cross-sectional study

**DOI:** 10.1101/2025.02.25.25322838

**Authors:** Enrique Rayo, Giulia Malingamba, Hanna Marti, Delia Onorini, Cory Leonard, Nicola Low, Benjamin Hampel, Nicole Borel

## Abstract

**Objective:** *Chlamydia trachomatis* (CT) and *Neisseria gonorrhoeae* (NG) are the most commonly reported sexually transmitted infections globally. Anorectal CT/NG detection among men who have sex with men (MSM) and co-infections are common. Epidemiological studies suggest that CT/NG co-infections might result in greater bacterial load and transmissibility than single infection. The purpose of this study was to compare bacterial load and symptoms between CT/NG single and co-infections in MSM.

**Methods:** MSM positive for CT or NG on a triple swab (throat, urethra, and rectal locations combined) were enrolled. Before treatment, they self-collected anorectal swabs. Bacterial loads for CT/NG were calculated using real-time PCR, and compared between single or co-infected individuals, and with or without rectal symptoms.

**Results:** We enrolled 382 MSM from December 2021 to December 2024. Among all samples: total CT (n=114), CT/NG co-infection 29/114 (25.4%); total NG (n=125), CT/NG co-infection 29/125 (23.2%). The bacterial loads in single and co-infected samples were comparable. The mean difference between CT alone and CT/NG was 0.40 copies/mL (95% CI: [-0.09, 0.11], P value = 0.107). The mean difference for NG alone and CT/NG was 0.24 copies/mL (95% CI: [- 0.49, 0.99], P value = 0.498). Among 382 MSM, 15.4% (n=59/382) experienced anorectal symptoms. There was no statistical difference in bacterial burdens between symptomatic and asymptomatic (CT difference of the means 0.52 copies/mL, 95% CI: [-0.51, 1.55]; P value = 0.313) (NG difference of the means 0.63, CI: [0.01, 1.28]; P value = 0.05).

**Conclusions:** In contrast to prior research, we found similar bacterial burdens in anorectal MSM samples with single CT/NG vs coinfection. Further research is needed to understand the clinical implications of CT/NG co-infections. Future should investigate factors influencing anorectal CT/NG bacterial burden, transmissibility, and susceptibility, including the function of PrEP and the rectal microbiota.

**Key Messages:**

- Epidemiological and modelling studies have generated the hypothesis that the bacterial load in co-infections of *Chlamydia trachomatis* (CT) or *Neisseria gonorrhoeae* (NG) may be higher than that of either organism alone.
- In this study, we found no statistical evidence of differences in the bacterial load of CT or NG between single and co-infections in men who have sex with men (MSM) using HIV pre-exposure prophylaxis, and no differences in bacterial loads between symptomatic and asymptomatic individuals.
- Further research is needed to understand factors affecting bacterial load, transmissibility and susceptibility to anorectal CT and NG infections, including the roles of organism viability and prophylactic drug use among MSM populations.

## INTRODUCTION

*Chlamydia trachomatis* (CT) and *Neisseria gonorrhoeae* (NG) are the most commonly reported bacterial sexually transmitted infections (STIs) globally [1]. Coinfections of both pathogens are common, with up to 40% of individuals with NG also having CT. Thus, a biological relationship between CT/NG has been hypothesized, as an increase in CT transmissibility or susceptibility would be required to duplicate reported levels of co-infection. Higher gonococcal loads have been found to increase transmission probability [2], so higher bacterial loads in CT/NG co-infections than single infections might result in more symptomatic infections and increase transmissibility. In a retrospective study from the Netherlands, NG bacterial loads were reported to be higher in CT/NG coinfection than in NG infection alone in anorectal swabs from men and in vaginal samples from women [3]. Bacterial loads did not differ between symptomatic and asymptomatic individuals. The study, however, did not report CT loads in single or co-infections.

Anorectal CT and NG alone and co-infections are generally common among men who have sex with men (MSM) [4,5]. Prevalence is even higher among the subset of MSM who take pre-exposure prophylaxis (PrEP) to prevent HIV infection [6], making them an important population among whom to investigate the epidemiology and clinical impact of CT/NG co-infections. The objectives of this study were to investigate CT and NG anorectal infections load among MSM using PrEP, to compare bacterial loads in single and co-infections and to compare bacterial loads in symptomatic and asymptomatic infections.

## METHODS

Participants in this research were part of the SwissPrEPared Study, at the Checkpoint Clinic Zurich, Switzerland, which provides PrEP and sexual health care for people at high risk of acquiring HIV infection (swissprepared.ch). SwissPrEPared participants attend follow-up visits at the clinic every 3 months, which include STI testing. Self-collected swabs from the urethra, anorectal canal and throat are placed in a single container for PCR detection of CT and NG. Those with a positive result for CT and/or NG who returned to the clinic were invited to enrol in our study before antimicrobial treatment. Study participants provided a self-collected anorectal swab, which was stored at 4°C, and provided information about anorectal symptoms. Staff from our laboratory collected all swabs 24 to 72 hours post-sampling, transported them to the laboratory (2.5km away) on ice and stored them at -80 °C. DNA was extracted from a 200 μL starting volume and eluted in 60 μL (DNeasy® Blood and Tissue Kit protocol, Qiagen; Germany). DNA extracts were tested for CT and NG using real-time PCR, as described previously [7]. Based on the amplification cycle threshold, bacterial loads were expressed as log10 copies/mL. We calculated co-infection proportions (with 95% confidence intervals, CI), mean bacterial load and standard deviation (SD) for single infections, co-infections and the difference between the means (with 95% CI). We compared the difference between the means for CT or NG infection alone and CT/NG co-infection, and for individuals with and without anorectal symptoms, using Tukey’s HSD (honestly significant difference) test in the R statistical software (R Core Team, 2020). All participants provided written informed consent. Ethical approval was granted by the Zurich cantonal ethics committee (project number 2021-01802).

## RESULTS

Between December 2021 and December 2024, we collected 485 anorectal swabs. We excluded non-PrEP users (55 MSM) and duplicates (47 samples from 42 different MSM, only first sample collected used), resulting in 382 unique PrEP anorectal swabs. Of these, 22.3% (n=85/382) were positive for only CT and 25.1% (n=96/382) were only positive for NG. Among 114 with CT infection, 29 (25.4%) were co-infected with NG. Among 125 with NG, 29 (23.2%) were co-infected with CT. In total, 7.6% (n=29/382) were positive for both CT and NG. In single infections, NG were higher than CT loads; difference between the means 0.67, CI: [0.25, 1.09], P value 0.0019). In co-infections, NG loads were higher than CT loads (difference of the means 0.82, CI: [0.05, 1.59], P value 0.037; significant at α=0.05). The CT load in CT/NG co-infection was similar to single infection CT loads (difference of the means 0.40 copies/mL, 95% CI: [-0.09, 0.89], P value 0.107). The NG load in CT/NG co-infection showed no significant difference compared to single infection NG loads (difference of the means 0.24 copies/mL, 95% CI: [-0.49, 0.99], P value 0.498) (Fig. 1).

**Figure 1:**
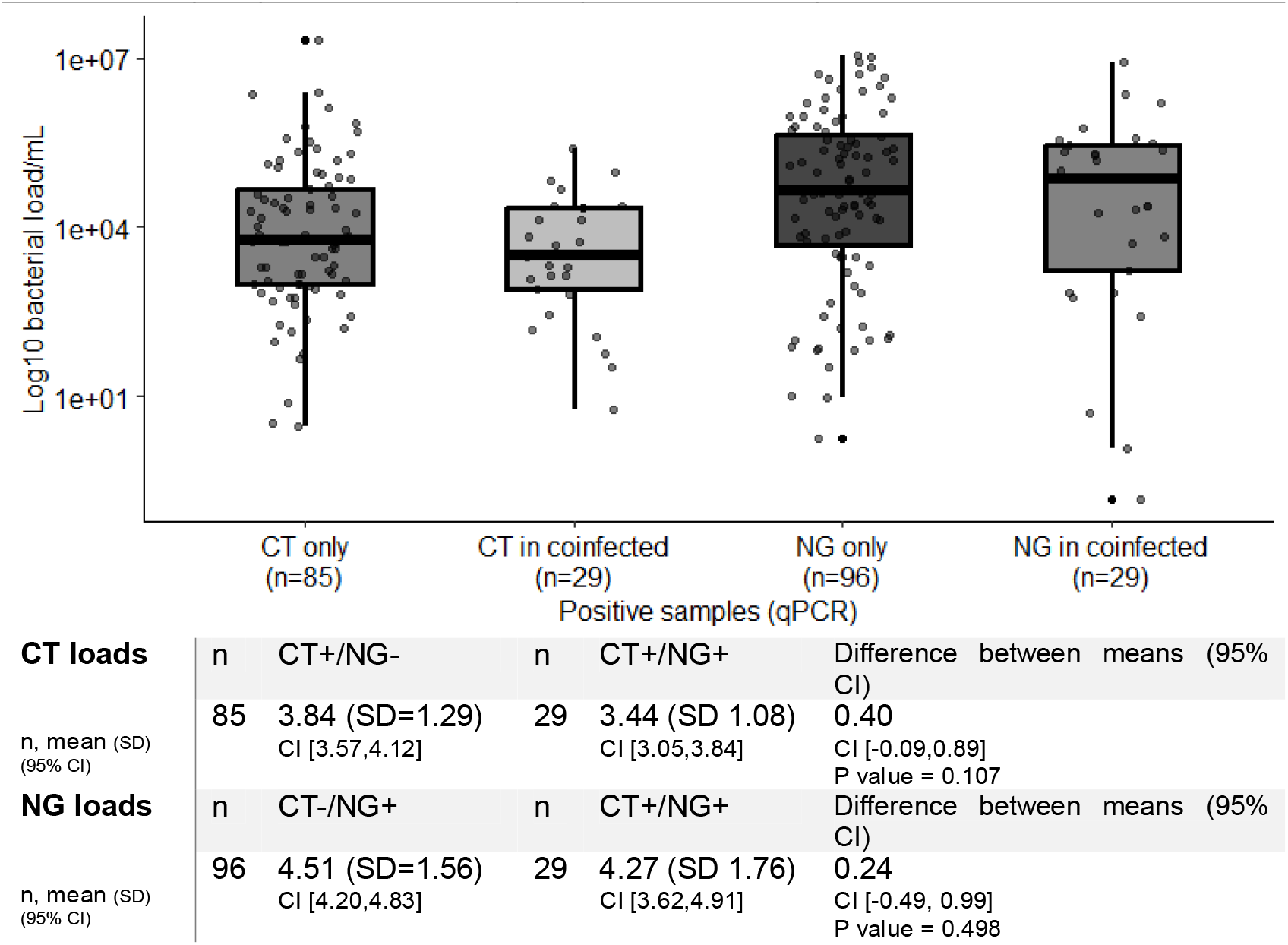
Box plot show bacterial loads assessed by real-time quantitative PCR assays for *C. trachomatis* and *N. gonorrhoeae* in samples with single and co-infection, and differences (co-infected minus single), tabulated below. The central line within each box represents the median, the top and bottom of each box mark the interquartile range (IQR) from the first to the third quartile, capturing the middle 50% of values. Whiskers extend up to 1.5 times the IQR from each quartile, and points beyond the whiskers represent outliers. CI, confidence interval; CT, *C. trachomatis*; NG, *N. gonorrhoeae*; SD, standard deviation.

Among 382 MSM, 15.4% (n=59/382) reported anorectal symptoms. Bacterial loads for CT did not differ between symptomatic and asymptomatic individuals (mean CT copies in symptomatic 4.09, SD 1.74; mean CT copies in asymptomatic 3.57, SD 1.29; differences between the means 0.52, CI: [-0.51, 1.55], P value 0.313). NG bacterial loads in symptomatic individuals tend to be higher than in asymptomatic individuals, but the difference does not reach statistical significance (mean NG copies in symptomatic 4.67, SD 1.59, mean NG copies in asymptomatic 4.04, SD 1.79; difference between the means 0.63, CI: [0.01, 1.28], P value 0.05,).

## DISCUSSION

In this cross-sectional study among MSM in the SwissPrEPared study, we did not find statistical evidence of a difference in bacterial loads in CT/NG anorectal co-infections compared with either CT or NG alone or in symptomatic compared with asymptomatic infections.

The strengths of our study include the structured study design, which allowed for the consistent collection of samples over time. The collection of samples prior to antibiotic treatment ensured that bacterial loads were all from untreated infections. Our study has some limitations. The sample size was relatively small, and we focused exclusively on anorectal infections among MSM on PrEP, which limits generalisability. The limited number of co-infections (n=29) reduces the precision of bacterial load estimates. However, we believe it is unlikely that we missed a clinically significant difference, as the differences between the means were minimal (Fig. 1).

Our results for NG loads in anorectal swabs in men differ from those reported by van Dessel et al., who found higher NG loads in CT/NG coinfected samples (mean 4.70 log_10_ copies/mL, 95% confidence interval 2.21-6.39, n=210) than in samples with NG alone (4.09, 1.57-6.64, n=825) [3]. In our study, the mean NG load in CT/NG co-infection was slightly lower than that in single infection (Fig. 1). The reason for the different findings might results from differences between the study participants in the two studies. Our study included only MSM taking PrEP for HIV prevention, where participants receive frequent healthcare visits. Frequent STI testing is likely to detect newly acquired infections, which might have higher bacterial loads; there might therefore be a smaller difference in bacterial load between single and co-infection. In the study by van Dessel et al. information about PrEP use and the time from infection to sampling was not reported. The van Dessel study also reported that there was no difference in a smaller sample of anorectal swabs from women. Mean load in co-infection (4.42, 2.18-6.50, n=35) was, however, higher than in single infection (3.81, 1.82-5.95, n=105). For both men and women, the difference between the means is 0.61 log_10_ copies/ml. Among women, the P value (0.13) provides weak statistical evidence and is consistent with the smaller sample size. In both studies, the presence or absence of symptoms was not associated with a difference in bacterial load.

Clinical relevance of CT/NG co-infections remains uncertain. Amongst those with NG, the proportion of anorectal CT/NG co-infections is not well-described but might be less common than vaginal co-infections. Amongst anorectal samples with NG detected, the proportions with both CT and NG are similar among men in our study (23%), and both men (20%) and women (25%) in the study by van Dessel et al. In vaginal samples from women in the study by van Dessel et al. 40% of samples with NG (n=349) also had CT detected (n=142). This proportion is consistent with other published studies of women with NG [8– 10]. Future studies should investigate CT/NG co-infections in a range of anatomical sites in both men and women and in those with both CT and NG. Factors of relevance include the role of medication including PrEP, the role of viability or non-viability of PCR-detected pathogens, and the potential role of other concomitant bacteria.

## Data Availability

All data produced in the present study are available upon reasonable request to the authors.

## References

1 Rowley J, Hoorn S Vander, Korenromp E, et al. Chlamydia, gonorrhoea, trichomoniasis and syphilis: global prevalence and incidence estimates, 2016. Bull World Health Organ. 2019;97. doi: 10.2471/BLT.18.228486

2 Stupiansky NW, Van Der Pol B, Williams JA, et al. The natural history of incident gonococcal infection in adolescent women. Sex Transm Dis. 2011;38:750–4. doi: 10.1097/OLQ.0B013E31820FF9A4

3 van Dessel HA, Dirks JAMC, van Loo IHM, et al. Higher Neisseria gonorrhoeae bacterial load in coinfections with Chlamydia trachomatis compared with Neisseria gonorrhoeae single infections does not lead to more symptoms. Sex Transm Infect. 2024;100:127–8. doi: 10.1136/SEXTRANS-2023-055977

4 Traeger MW, Cornelisse VJ, Asselin J, et al. Association of HIV Preexposure Prophylaxis With Incidence of Sexually Transmitted Infections Among Individuals at High Risk of HIV Infection. JAMA. 2019;321:1380–90. doi: 10.1001/JAMA.2019.2947

5 Khosropour CM, Coomes DM, Leclair A, et al. High Prevalence of Rectal Chlamydia and Gonorrhea Among Men Who Have Sex With Men Who Do Not Engage in Receptive Anal Sex. Sex Transm Dis. 2023;50:404–9. doi: 10.1097/OLQ.0000000000001803

6 Jongen VW, Van Der Loeff MFS, Van Den Elshout M, et al. Bacterial sexually transmitted infections are concentrated in subpopulations of men who have sex with men using HIV pre-exposure prophylaxis. AIDS. 2023;37:2059–68. doi: 10.1097/QAD.0000000000003676

7 Onorini D, Leonard CA, Phillips Campbell R, et al. Neisseria gonorrhoeae Coinfection during Chlamydia muridarum Genital Latency Does Not Modulate Murine Vaginal Bacterial Shedding. Microbiol Spectr. 2023;11. doi: 10.1128/SPECTRUM.04500-22

8 Althaus CL, Turner KME, Schmid B V., et al. Transmission of Chlamydia trachomatis through sexual partnerships: a comparison between three individual-based models and empirical data. J R Soc Interface. 2012;9:136–46. doi: 10.1098/RSIF.2011.0131

9 Forward KR, Frcpc MD. Risk of Coinfection with Chlamydia trachomatis and Neisseria Gonorrhoeae in Nova Scotia. Canadian Journal of Infectious Diseases and Medical Microbiology. 2010;21:e84–6. doi: 10.1155/2010/760218

10 Seo Y, Choi KH, Lee G. Characterization and Trend of Co-Infection with Neisseria gonorrhoeae and Chlamydia trachomatis from the Korean National Infectious Diseases Surveillance Database. World J Mens Health. 2021;39:107– 15. doi: 10.5534/WJMH.190116

